# Psychosocial factors contributing to a syndemic of substance use and HIV risk among Floridian transgender women

**DOI:** 10.1101/2024.10.04.24314900

**Authors:** Daniell Sullivan, Rachel Clarke, Hannah Wilson, Jessy Dévieux, Elena Cyrus

## Abstract

**Background:** Transgender women are at heightened risk of HIV and substance misuse due to systemic discrimination and structural healthcare barriers at the macro, meso, and micro levels. Floridian Transgender Women (FTW) face unique challenges in an environment with limited resources and inconsistent epidemiological data. Syndemic theory, which examines how co-occurring conditions like HIV and substance use disorders (SUD) are driven by large-scale factors, has not been fully explored in FTW.

**Objective:** This study aimed to identify psychosocial factors contributing to substance use and HIV risk from a syndemic perspective among FTW.

**Methods:** From May 2021 to November 2023, over 500 at-risk FTW were recruited from community partner agencies in Miami-Dade and Orlando counties for a larger prospective study. Univariate analysis identified psychosocial factors, while bivariate analysis explored their contributions to the syndemic of substance misuse and HIV risk.

**Results:** Out of 160 participants screened, 55.6% were ineligible, leaving 89 enrolled. Participants ranged in age from 18 to 63, with 25% identifying as people of color. Mental illness was reported by 62.9%, with depression and anxiety being the most common diagnoses. However, only 29.2% had accessed mental health care in the past year. Despite high rates of substance use—48.8% reported drug use during sex, 46.1% likely had an SUD, and 25.8% reported hazardous drinking—68.9% perceived their quality of life as high. Drug use during sex was significantly associated with intimate partner violence (IPV) (p=.01) and perceived quality of life (p=.003).

**Conclusion:** FTW face significant challenges, including discrimination, mental illness, trauma, and IPV, which increase the risk of substance use, particularly marijuana and alcohol. Despite these challenges, many maintain a high perceived quality of life. The syndemic interaction calls for comprehensive interventions addressing financial, mental health, and accessibility barriers. Further research into socio-political stressors that exacerbate substance use and high-risk behaviors is recommended.

## Background

Nationally, transgender women are disproportionately susceptible to HIV due to significant healthcare barriers, with an estimated prevalence of 14.1% in some surveillance groups compared to their cisgender counterparts’ 0.5%.^1^ Additionally, this population also engages in higher rates of illicit substance use, attributed to various psychosocial factors such as housing instability, mental illness, and socioeconomic status. Substance use in transgender women is known to drive high risk sexual behaviors (HRSBs) and thereby HIV infection risk.^2,3,4,5^ In other words, this community exhibits an HIV and substance use syndemic, a phenomenon where societal circumstances affect how two or more health conditions interact within a population. ^6^

Importantly, Florida (FL) has the second largest population of trans adults in the United States, mostly concentrated in Orlando and Miami, so the proportion of Floridian transgender women (FTW) is somewhat reflective of the national community.^7,8^ Limited substance use and HIV epidemiological data exists for FTW. In fact, to our knowledge, there are no consistent estimates for FTW HIV prevalence. This is a harrowing realization since FL leads the nation in new HIV cases.^9^ Previously, we estimated the FTW HIV prevalence to be around 38.6%, nearly triple the national transgender women prevalence and 64 times greater than the prevalence of Florida’s general population.^9,10,11,12^ Moreover, FL has the highest rate of new HIV cases in the nation, partially caused by a universal lack of access to care.^9,13^ Therefore, we sought to identify certain psychosocial factors that syndemically influence this disproportionality in HIV prevalence and illicit substance use rates utilizing Bronfrenbrenner’s ecological systems theory.

At the foundational level, this theory examines how bioecological interactions on varying scales influence an individual’s development. These scales include microsystems, consisting of direct interpersonal relationships; mesosystems comprised of multiple microsystems; macrosystems, such as socioeconomic status and cultural ideologies. ^14^ When applied to populations, the theory appropriately conveys the interconnectedness of psychosocial factors that contribute to a syndemic. For example, one study employed this approach to identify psychosocial factors that synergistically influence HIV risk in black transgender women and men who have sex with men.^15^ However, no study to our knowledge has applied this framework to FTW specifically.

Because of the regional sociopolitical climate, FTW are understudied and research directed towards this population is underfunded. Therefore, FTW are especially vulnerable compared to their national counterparts, considering FL has enacted six anti-LGBTQ laws in the past year, significantly limiting FTW’s access to treatment.^16^ Moreover, state legislation that restricts TW’s access to gender-affirming care may promote medical mistrust that prevents TW from seeking care elsewhere, further contributing to the high HIV prevalence in this population.^16^ For the above reasons, the present study focused on this specific geographic population. The main objective of this study was to identify and explore associations between psychosocial factors that contribute to illicit substance use and HIV infection risk from a syndemic perspective in FTW.

## Methods

### Participants

FTW were recruited throughout central and southern Florida between May 2021 to November 2023 as part of a longitudinal parent study termed To Reach Unrestricted Services for Transwomen (TRUST) (PI: Dr. Elena Cyrus). Recruitment involved partnering with Florida federally qualified healthcare centers (FQHC) in the Orlando and Miami areas serving transgender women,, as well as social media outreach. The Miami centers included “Sunserve” and “Arianna’s Center,” in Miami-Dade County and “26 Health” in the Orlando area. Individuals who were assigned male sex at birth and currently identify as adult (≥18years) female were enrolled in the study. Participant HIV serostatus was collected via self-report and laboratory confirmatory testing. Those <18 years old or HIV positive were excluded from the study. Out of 160 participants screened, 90 met study criteria and completed the baseline questionnaire at the time of analysis. At baseline, each participant completed an interviewer-administered questionnaire using Research Electronic Data Capture (REDCap) and included information related to participant demographics and behaviors.^17,18^ REDCap is a secure, web-based software platform designed to support data capture for research studies, providing 1) an intuitive interface for validated data capture; 2) audit trails for tracking data manipulation and export procedures; 3) automated export procedures for seamless data downloads to common statistical packages; and 4) procedures for data integration and interoperability with external sources.

### Ethics Statement

Because this was secondary analysis of an existing dataset, this research project received IRB exemption from the UCF IRB, as all data used for the study and subsequent analyses had already been de-identified, and the original study was previously approved for conduct of human subjects research by same IRB

### Variables

#### Demographics

Participants provided information related to race, ethnicity, sexual orientation, monthly income, and age.

#### HIV Infection Risk

HRSBs are highly predictive of HIV infection risk^19^ and were therefore used as a proxy variable for measurable HIV infection risk. HRSBs included unprotected sex, or engaging in condomless penetrative/receptive anal sex within the last 6 months; multiple sex partners, or having ≥2 sex partners in the past 6 months; drug use during sex, or using drugs immediately before or during a sexual encounter in the last 6 months; and alcohol use during sex, or using alcohol immediately before or during a sexual encounter in the last 6 months.

#### Substance Use

Participant substance use data included substance type and frequency, ranging from daily to one time in the last 6 months. Substance types were then classified according to the following Drug Recognition Expert categories: cannabinoids included marijuana and synthetic cannabis, hallucinogens included LSD and MDMA, inhalants included amyl nitrite (i.e. poppers), stimulants included cocaine and methamphetamine, depressants included benzodiazepines and GHB, narcotic analgesics included opioids, and dissociative anesthetics included ketamine.^20^ Hazardous drinking was defined by having an Alcohol Use Disorder Identification Test (AUDIT) score ≥8, which correlates to a higher likelihood of having an alcohol use disorder and consequently requires further intervention.^21^ Substance use disorders (SUD) likely were defined as having a Drug Abuse Screening Test – 10 (DAST-10)^22^ score of ≥3, which has a high likelihood of meeting DSM criteria for an SUD.^23^

#### Psychosocial Factors

Discrimination was defined as having a prior subjective experience of physical or verbal discrimination based on gender identity/presentation, race, ethnicity, or skin color. Trauma was defined as having history of experiencing at least one traumatic event during this lifetime. Intimate partner violence (IPV) was defined as having a history of experiencing sexual or physical violence at the hands of a partner. Mental health history referred to ever having a history of receiving a mental illness diagnosis (yes or no). If yes, current mental illness was subclassified based on diagnosis, including depression, anxiety, and PTSD. Mental health care discrimination was defined as having experienced problems seeking mental health care due to gender presentation. Participants were also asked on whether or not they accessed mental health care resources in the past 12 months. Homelessness was defined as living without housing at any point in time. Incarceration was defined as being imprisoned at any point in time. Perceived quality of life (pQoL) was measured on a categorical scale from 0-4, where 0 indicated very poor quality of life and 4 reflected a very high quality of life. Those who answered 0-1 were categorized as having a poor pQoL. Those who answered 2, which equated to neither poor nor high quality of life, were categorized as having an average pQoL. Participants who answered 3-4 were categorized as having a high pQoL.

### Data Analysis

Income and age were open ended and continuous. Nominal categorical variables included current mental illness, substance type, and race. pQoL was categorized as the only ordinal variable as previously described. All other variables were binary. Univariate and bivariate frequencies were conducted to describe the study population, psychosocial factors, and risk behaviors. Variable frequencies are reported based on the valid percentage of responders. Chi Square tests were conducted to examine associations between psychosocial factors and HRSBs and substance yse, where associations were considered significant at alpha = 0.05. Statistical Package for the Social Sciences (SPSS) version 28 software was utilized to conduct all analyses.

## Results

### Descriptive Results

Participant demographic characteristics are presented in Table 1. The mean age was 28.4 years (standard deviation = 8.8, range = 18.0-63.42). Within our sample, 38.6% of participants identified as Hispanic. Overall, 54.5% of participants identified as White, 13.6% as African American, 2.3% as Asian, and 13.6% as Multiracial. Reported sexual orientation was broad, but the majority of participants identified as either pansexual or queer (36.3% total). On average, participants only had a monthly income of $1785.90 (standard deviation = $1634.7, range = $0-8000).

**Table 1.**
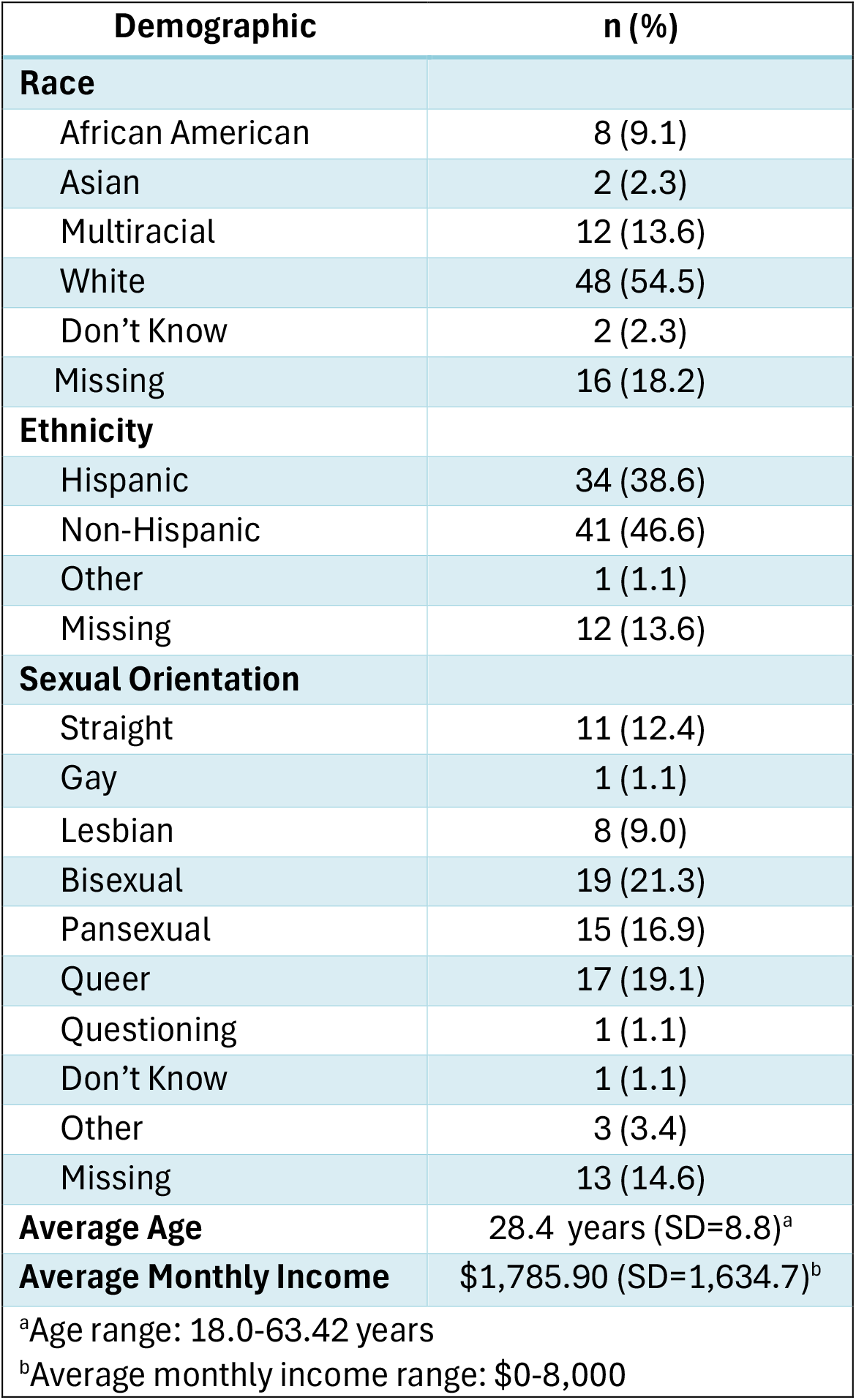
Participant demographics for the TRUST Study (n=89).

Psychosocial factors are show in Table 2. The majority of particpants reported experiencing discrimination, trauma, and mental illness at one point in their lifetime at 88.8%, 87.6%, and 62.9%, respectively; yet, 69.3% of these women reported having a high pQoL. Of those previously diagnosed with a mental health condition, the majority experienced comorbid illness, particularly anxiety and depression as shown in Table 3.

**Table 2.** Cross-Sectional self-reported psychosocial factors for the TRUST Study at the Baseline Visit (n=89).

| **Psychosocial Factors** | **%** |
| --- | --- |
| Diagnosed with a mental illness | 62.9 |
| Mental health care discrimination in past 12 mths | 14.6 |
| Accessed mental health services in past 12 mths | 29.2 |
| Ever incarcerated | 6.7 |
| Ever experienced a traumatic event | 87.6 |
| Ever experienced homelessness | 29.2 |
| Ever experienced discrimination | 88.8 |
| Ever experienced intimate partner violence | 43.8 |
| High perceived QoL | 68.5 |
| Low perceived QoL | 9.0 |
| Hazardous drinking (AUDIT $\geq 8$) | 25.8 |
| SUD likely (DAST $\geq 3$) | 46.1 |

**Table 3.** Participants’ current mental health status among TRUST baseline participants who reported ever having a mental health diagosis (n=56).

| **Mental Health Diagnosis** | **n (%)** |
| --- | --- |
| Depression | 4 (7.1) |
| PTSD | 6 (10.7) |
| Anxiety | 9 (16.1) |
| Depression + PTSD | 1 (1.8) |
| Depression + Anxiety | 22 (39.2) |
| PTSD + Anxiety | 1 (1.8) |
| Depression + PTSD + Anxiety | 13 (23.2) |

Table 4 shows the rates of HRSBs, as 42.0% had multiple sex partners in the last 6 months, and nearly 18.2% engaged in unprotected sex. 35.2% used alcohol before sex, and 26.1% were considered to engage in hazardous levels of drinking. 45.5% report using drugs during sex, and 46.6% were considered to likely have an SUD.

**Table 4.** Self-reported high risk sexual behaviors among TRUST Baseline partipants who were sexually active Study (n=84).

| **High Risk Sexual Behaviors among sexually active participants** | **n (%)** |
| --- | --- |
| ≥2 sex partners in the last 6 months | 37 (44.0) |
| **Missing** | 20 (21.3) |
| Alcohol use during sex in the last 6 months | 32 (38.1) |
| **Missing** | 20 (21.3) |
| Drug use during sex in the last 6 months | 41 (48.8) |
| **Missing** | 20 (21.3) |
| Condomless sex in the last 6 months | 16 (19.0) |
| **Missing** | 62 (73.8) |

Table 5 shows the percentage of participant substance use classified into individual substances. Participants were most likely to use cannabinoids with 73.0% participants using over the last 6 months. 51.1% of participants reported using marijuana over the last month and 100% of responders reported using at least once in the past 6 months, as the remaining 27% did not provide input to their marijuana use behaviors. The next most used drugs over the past 6 months were hallucinogens at 36.4%, inhalants at 21.6%, and stimulants at 14.8%.

**Table 5.**
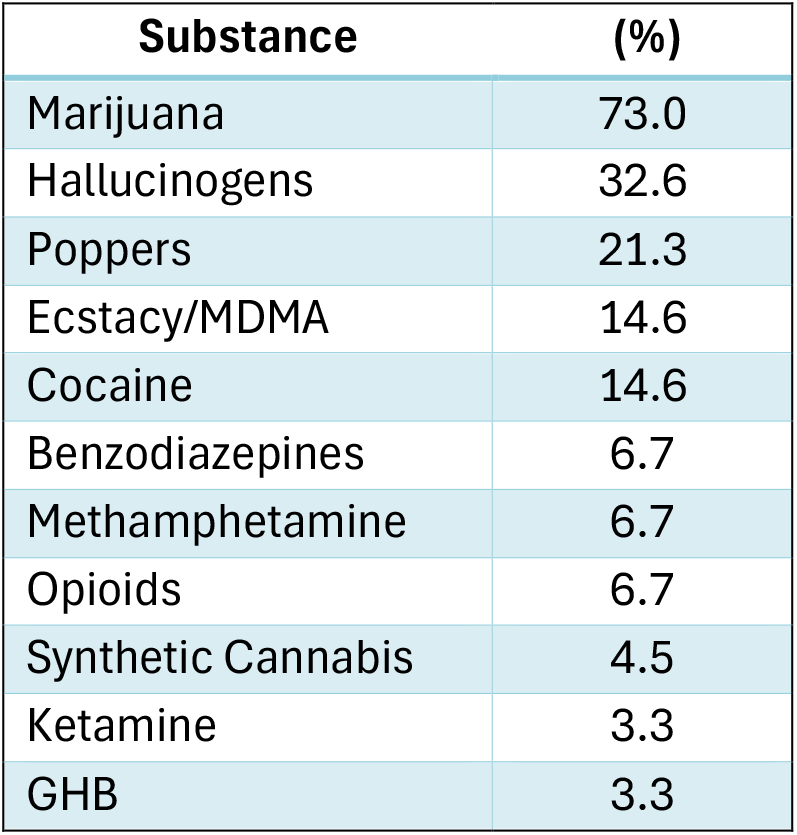
Participant substance use in the past 6 months reported by baseline TRUST participants (n=89).

### Analytic Results

When examining the relationships between psychosocial factors with HRSBs and substance use, chi squared analyses revealed a significant association between drug use during sex and IPV (p=.017) as well as pQoL and Drug use during sex (p = 0.003). All other associations between SPFs and HRSBs were nonsignificant. Secondary analysis between HRSBs revealed a significant correlation between alcohol use during sex and drug use during sex (p<.001).

## Discussion

We investigated the influence of various psychosocial factors on the syndemic of HIV and substance use disorder (SUD) among Floridian transwomen. Our analyses reveal multiple syndemics between psychosocial factors and high-risk sexual behaviors (HRSBs), as well as within these categories themselves. Participants experienced disproportionately high rates of lifetime homelessness, more than double the national average.^24^ Additionally, 62.9% of participants had been diagnosed with a mental health condition, a significant increase compared to the 22.8% national average for adults diagnosed with mental illness in the past year.^25^ This high prevalence may be explained by the almost universal reporting of trauma and discrimination. Comorbid depression and anxiety were the most commonly reported conditions, with 44.9% of participants diagnosed with depression—slightly higher than the estimated 31% in the national transgender population, potentially influenced by state-specific anti-transgender legislation.^26^

Despite elevated rates of mental illness, only 29.2% of participants had accessed mental health care in the past year, representing about one-third of those with a diagnosed condition. This figure contrasts with the 59.8% of the general population who accessed mental health care in 2023, highlighting significant barriers for transwomen, such as medical mistrust, a lack of mental health providers, and limited socioeconomic resources.^25^

Substance use was also notably high in this cohort. One in four participants reported hazardous drinking, as indicated by their AUDIT scores—higher than the 18% rate reported among transgender individuals nationally.^27^ This suggests that Floridian transwomen may be at even greater risk for alcohol use disorders (AUDs) than their peers in other states. All participants who responded about illicit substance use had used marijuana at least once in the past six months, and over 50% used it at least monthly—nearly triple the national average for individuals aged 12 and older.^25^ For comparison, past three-month cannabis use among U.S. transgender individuals was previously reported at 24.4%, further suggesting a potential geographic influence.^28^ Additionally, participants exhibited significantly higher use rates of other illicit substances, particularly hallucinogens, cocaine, and MDMA. Nearly half (46.1%) of participants were likely to have a substance use disorder (SUD), more than double the rate seen in the general population.^25^ This widespread recreational drug use correlates with the 48.8% of participants who reported drug use during sex, a known HRSB and predictor of HIV risk.

Interestingly, higher rates of intimate partner violence (IPV) were associated with drug use during sex. This may reflect a coping mechanism related to transactional sex or pervasive violence experienced by the population. Transwomen are already at an increased risk for IPV, and those with lower socioeconomic status—evidenced by a median income of just under $1,800—are especially vulnerable.^29^ Partner abuse, particularly when sexual in nature, can trigger a trauma response in subsequent sexual activities, potentially leading to self-medication to suppress this response.^30^

Further analysis revealed a significant association between two HRSBs: drug use during sex and alcohol use during sex, a relationship well-established in existing literature. Concurrent substance use may synergistically increase Floridian transwomen’s HIV infection risk compared to the use of a single substance.^31^ These within-group associations between psychosocial factors and HRSBs exemplify the classic characteristics of a syndemic: specific psychosocial variables have a combinatorial effect on behaviors, health outcomes, and subsequent HIV risk. In other words, the factors driving the HIV disease burden are additive, not independent.^6^ Low income and high levels of discrimination—both seen in this study—likely prevent transgender women from accessing health care and resources, contributing to high HIV prevalence.^32^ Additionally, a lack of educational resources tailored to this specific community further lowers HIV risk perception and increases transmission rates.^33^

Despite facing disproportionately high levels of trauma, IPV, discrimination, mental illness, and financial adversity compared to cisgender individuals, Floridian transwomen report a surprisingly high perceived quality of life (pQoL), showcasing the resilience of this community.^7,34,35^ The relationship between high pQoL and drug use during sex observed in this study may be bidirectional. Some transwomen may use drugs as a coping mechanism that temporarily enhances their perceived QoL, while also impairing their awareness of the consequences of HRSBs.^10^ On the other hand, many transwomen engage in sex work for financial stability, where they are more likely to be exposed to and coerced into drug use..^36^

Several limitations of this study should be noted. The sample size was underpowered, as the parent study was exploratory in nature, meaning only associations could be analyzed. Although most participants provided data for each variable, response rates varied between questions, reducing the study’s power. Notably, 74% of participants did not respond to questions about condom use, likely due to social desirability bias—the tendency to underreport socially undesirable behaviors such as sexual practices, drug use, and mental health conditions, while overreporting more acceptable behaviors.^37^ Moreover, the majority of participants identified as white, limiting the generalizability of these findings. Black transwomen were particularly underrepresented, possibly due to the chain referral recruitment method used in this study. A larger and more diverse sample size may yield more significant associations between psychosocial factors and HRSBs.

In the future, Northern Floridian transwomen should also be included in the sample to ensure the results are representative statewide. As the parent study (ENTRUST) progresses, participants’ HIV status will be reassessed to determine how these psychosocial factors and behaviors contribute to HIV infection risk. Data from those who become HIV-positive can be used to create predictive models for HIV risk in transgender women based on these factors and behaviors. Ultimately, targeted interventions addressing psychosocial factors—particularly violence and financial insecurity—can help reduce substance use rates and the HRSBs that drive HIV infection risk.

## Data Availability

All data produced in the present study are available upon reasonable request to the authors.

